# A Dichotomous Pain Scale Reduces Post-Operative Opioid Consumption in Outpatient ACL Surgery: Results from a Prospective Randomized Controlled Study

**DOI:** 10.1101/2025.09.22.25336386

**Authors:** Danielle M. Mullis, Joseph P. Donahue, Seth L. Sherman, Marc R. Safran, Geoffrey D. Abrams, Todd F. Alamin

## Abstract

**Background/ Aim:** This study is a prospective, randomized controlled clinical study investigating the effect of using a dichotomous pain scale on post-operative opioid use in patients undergoing anterior cruciate ligament (ACL) reconstruction compared with the standard of using a 10-pt VAS scale. The primary objective was to determine whether the use of a dichotomous pain scale would reduce the consumption of opioids within the first two weeks after surgery.

**Methods:** Twenty-nine patients undergoing outpatient ACL reconstruction were randomly assigned to either the Binary (n=14) or Scale (n=15) group and completed a survey every day for two weeks after surgery. The Binary group was asked whether their pain was “tolerable” or “intolerable.” The Scale group was asked to rate their pain on a scale from zero to ten, as per standard of care.

**Results:** Patients in the Scale group on average consumed approximately twice the number of morphine milligram equivalents per day than the Binary group (*p=*0.03). Two weeks after surgery, there was no significant difference between patient satisfaction nor average knee range of motion between groups (*p*=0.63, *p=*0.08).

**Conclusions:** The results from this study suggest that using a dichotomous tolerability-based pain scale instead of the NPRS could serve as one strategy to reduce opioid use in the post-operative period.

## Introduction

Anterior cruciate ligament (ACL) reconstruction is a very common orthopedic surgery mostly performed in the outpatient setting [1]. Despite how commonly this procedure is performed, it has been found to be one of the most painful orthopedic surgeries [2]. Historically, opioids are commonly used to manage post-operative pain, and there has been a shift towards using supplementary multimodal pain control regimens [3]. However, there are currently no standardized regimens for controlling post-operative pain [4-5].

The current opioid epidemic in the United States is a pressing public health crisis. Orthopedic surgeries, such as ACL reconstruction, continue to contribute to this issue. A study in 2014 found that orthopedists are one of the most likely specialists to prescribe opioids [6], and recent studies have found that opioids are still used by many patients after undergoing ACL reconstruction [7-9].

Opioids are associated with many risks, especially the development of opioid use disorder and subsequent overdose fatalities [10]. There is also risk for family members and individuals in contact with patients who are prescribed the opioids [10]. A shift in postoperative pain management is needed to combat the current opioid epidemic. Various publications have investigated the use of multimodal pain regimens [11-13], which is one promising strategy to reduce opioid consumption.

Another strategy to reduce the use of opioids and/or pain medications in the post-operative period is to better understand patients’ perception of pain or adjust expectations of pain management. A recent study [14] on patients with chronic pain looked at the effect of using a standardized pain tolerability question (“is your pain tolerable?”) in comparison to the traditional numeric pain rating scale (NPRS) (“rate your pain on a scale from 0 to 10”). Analyzing surveys of over 500 patients, they found that although the pain rating of “intolerable” is typically associated with a higher NPRS score, almost half the patients in the severe range of the NPRS (7-10) rated their pain as tolerable. The NPRS has been theorized to set unrealistic expectations for patients, as patients may understand the goal in pain management to achieve a “0” on the pain scale [15-16], which may lead to overmedication. It is interesting to note that one of the many policy changes in the mid-1990’s prior to the opiate epidemic was the codification of the NPRS as a “fifth vital sign” [17-18], to be used for all patient encounters.

This prospective randomized clinical trial investigates whether the use of a dichotomous tolerance-based pain scale from the initial interaction in the preoperative and then postoperative unit through follow-up reduces opioid consumption for patients in the first two weeks following ACL reconstruction in comparison to patients using the NPRS. We hypothesized that patients asked whether their pain was tolerable/intolerable would consume fewer morphine milligram equivalents (MME) in comparison to patients rating their pain on a scale from 0 to 10.

## Methods

Institutional Review Board approval was obtained prior to starting this study (Protocol 45010, approved on the 19^th^ of April 2018). Study participants were recruited between 05/30/2019 and 08/30/2021, and all participants provided informed written consent. The trial was registered with clinicaltrial.gov (NCT07065266). Thirty-six patients were approached about participating in this study. All patients were undergoing ACL reconstruction at one outpatient center. All participants voluntarily completed the research consent form. Patients were randomly assigned to either the Binary or Scale group; they were randomized by the Sports Medicine investigators prior to surgery, and each patient had a 50% chance of being assigned to either group.

The inclusion criteria was as follows: all adult patients who received elective outpatient ACL reconstruction at the outpatient location, with autograft or allograft. The exclusion criteria was as follows: pregnant women, any prior surgery to the operated knee, known allergies to opioid medication, any patients receiving extra-routing pain care (as compared with the average ACL patient at our institution) given by anesthesiologists, such as postoperative epidural analgesia or ultrasound-guided femoral nerve block. We aimed to enroll approximately 30 patients in the study, and we were unable to perform power analyses to determine desired sample size due to the lack of any prior literature on this topic.

The setting of the clinical trial was an outpatient surgical center with six operating rooms. The pre- and post-operative nursing staff, along with the anesthesia team at the surgical center, were consulted prior to the start of the trial to determine best methods of ensuring compliance with the nursing and postoperative medication order protocols as well as compliance with the study protocol. Computerized order sets were developed and incorporated into the electronic medical record. At the start of the trial, and intermittently throughout the trial, presentations were made to the nursing staff to ensure awareness of the trial. Each patient was identified as being in the experimental arm of the trial by an easily identifiable chart folder color to minimize the chance of inadvertent questions about NPRS.

For the subsequent fourteen days after the day of surgery, patients were asked what type and amount of the pain medications they used. They were specifically asked how much hydrocodone, oxycodone, and tramadol they were using on each post-operative day. MD Calc’s Morphine Milligram Equivalents (MME) Calculator [19] was used to calculate an MME consumed for all patients for each post-operative day. Participants were also asked to list any non-steroidal anti-inflammatory medications as well as any other pain medications they took that day for pain. The Scale group was asked to rate their current knee pain from a scale from 0 to 10. When patients were asked to rate their pain on a scale from 0 to 10, they were provided the following information: 0-1 no pain, 2-3 mild pain, 4-6 moderate pain, 7-8 severe pain, 9-10 worst pain imaginable. In the Binary group, patients were asked to select whether their current knee pain was either tolerable or intolerable.

On postoperative day 14, patients were also asked whether they had any of the following symptoms before their first follow-up visit: nausea, itching, decreased appetite, drowsiness, urinary retention, headache, and none. Data was recorded on the degree of knee flexion for each patient at their visit approximately two weeks after surgery. All participants, in both the Scale and Binary groups, were asked to rank their satisfaction with their pain management after surgery on a scale from 0 to 10, with 0 being the least satisfied and 10 being the most satisfied.

All statistical analysis was carried out using JMP (Cary, NC, USA). Average means were compared using two-tailed independent t-tests. Categorical variables were analyzed with Chi-squared tests. Linear regression analysis was carried out using Excel. Errors reported are standard deviations. All *p*-values less than 0.05 were considered statistically significant.

## Results

Thirty-six patients were approached about participating in this study. Twenty-nine patients consented to participate and filled in the survey for all 14 postoperative days (81%) (**Fig 1**). All other patients were excluded from the cohort. Out of the 29 total patients included in this study, 15 (52%) were randomly assigned the Scale group and 14 (48%) were assigned to the Binary group. In the Scale Group, 60% of participants were male and 40% were female, with an average age of 30 ± 8 years old. In the Binary group, 57% of participants were male and 43% were female, with an average age of 29 ± 4 years old. There was no significant difference between the distribution of patients who received an allograft versus autograft between groups; 11 of 15 (73%) received an autograft in the Scale group, and 13 of 14 (93%) received an autograft in the Binary group (*p*=.16, **Supplemental Table 1**).

**Figure 1:**
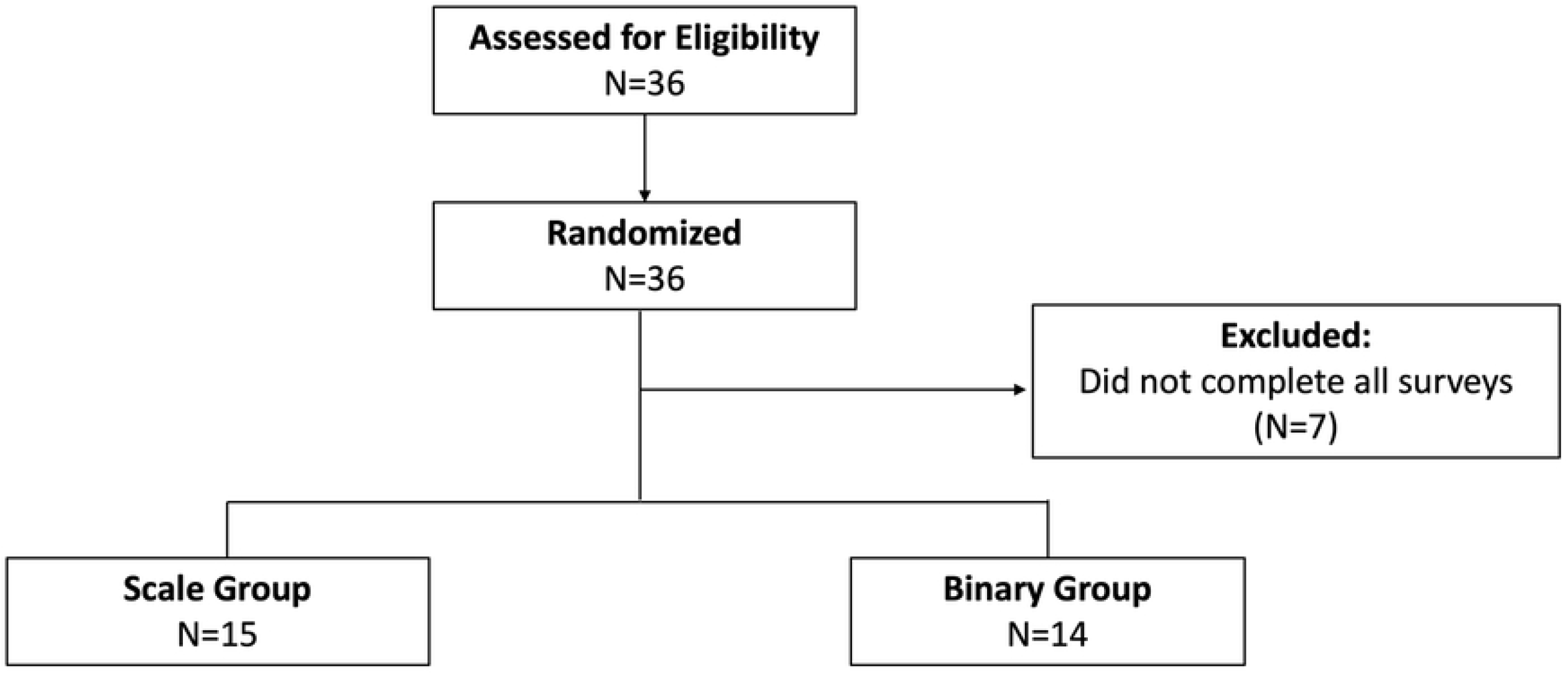
Flow Chart. Thirty-six patients gave voluntary consent to participate in this study. Seven patients did not complete all surveys on all fourteen post-operative days and were thus excluded from the final study cohort. Out of the 29 patients in the final cohort, 15 had been randomly assigned to the Scale group and 14 had been randomly assigned to the Binary group.

The Scale Group reported an average NPRS pain rating of 5 ± 2 on post-operative day (POD) 1, which decreased to 3 ± 2 by POD 14 (**Fig 2A**). In the Binary group, one person reported intolerable pain from POD 1 through POD 4, but between POD 5 and POD 14, every participant reported tolerable pain and no patients reported intolerable pain (**Fig 2B**).

**Figure 2:**
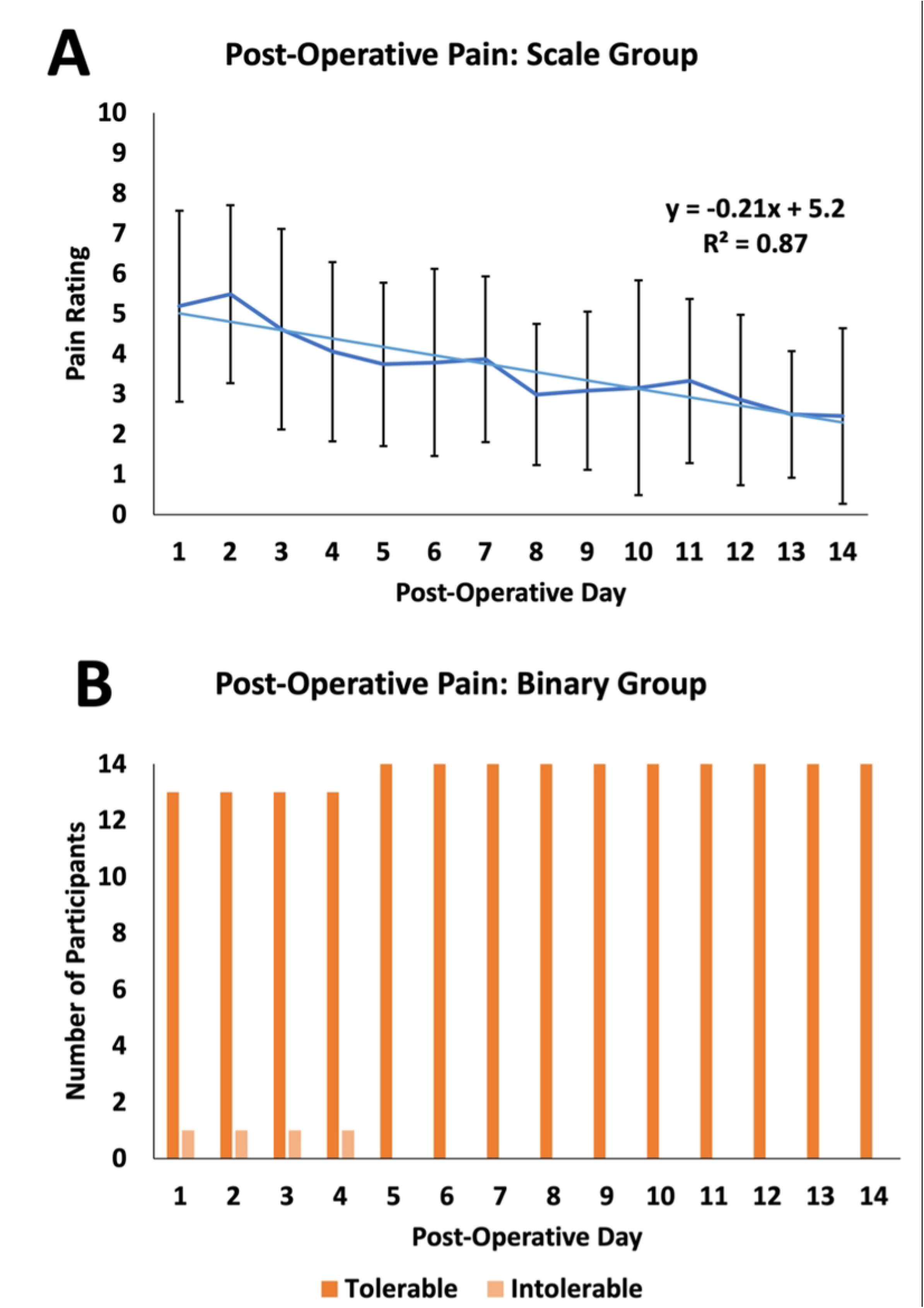
Post-Operative Pain Ratings Each Day. **A**) Post-operative pain scores for the Scale group. Pain scores decrease over time (y=-0.21x+5.2, R^2^=0.87). Error bars represent standard deviations. **B**) Post-operative pain scores for the Binary group. One patient rated their pain as “intolerable” for the first 4 days, and 100% of patients rated their pain as “tolerable” from post-operative day 5 until 14.

The experimental Binary group used significantly less total opiate pain medications per day in the two postoperative weeks by MME than the control Scale group (6.6 vs 13.4, *p*=0.03, **Fig 3 and Supplemental Table 3)**. To understand what this means clinically, one 5mg tablet of oxycodone is equivalent to 7.5 MMEs; our data shows that on average for all 14 post-operative day if patients were only taking oxycodone, each patient in the Binary group would have consumed approximately one 5mg pill of oxycodone per day, but the Scale group would have consumed double that amount. In the two weeks following surgery, patients in the Scale group used any type of pain medication for 12.3 ± 2.8 days in comparison to 10.4 ± 4.4 days in the Binary group (*p=*0.17, **Fig 4 and Supplemental Table 2**). Patients in the Scale Group used any opioid medication for 5.5 ± 3.8 days, and patients in the Binary Group used opioid medication for 4.6 ± 3.7 days. For the Scale group, the last post-operative day using opioids was POD 6; for the Binary group, the last post-operative day using opioid was POD 4 (*p*=0.21). Patients in the Scale group appeared to use acetaminophen and ibuprofen for more days after surgery (6.5 and days, respectively) than the Binary Group (4.8 and 1.6 days, respectively), although this did not reach statistical significance (*p*=0.31 and 0.19, respectively).

**Figure 3:**
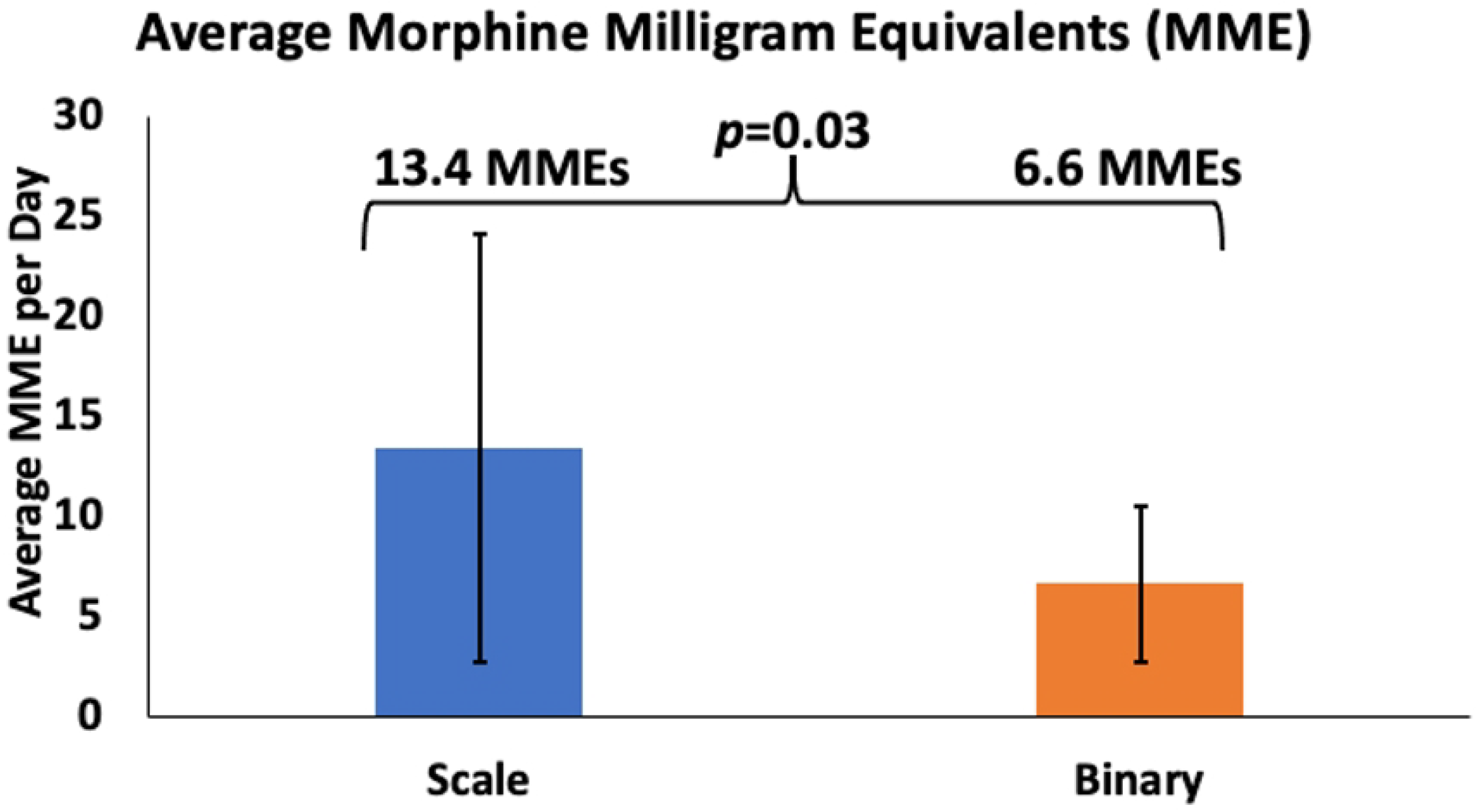
Average Doses of Opioids Per Day in the Post-Operative Period. **A**) Average morphine milligram equivalents (MME) per day were calculated for each patient. There was a significant difference between the Scale and Binary groups (13.4 vs 6.6 MMEs per day, *p*=0.03). Error bars represent standard deviations.

**Figure 4:**
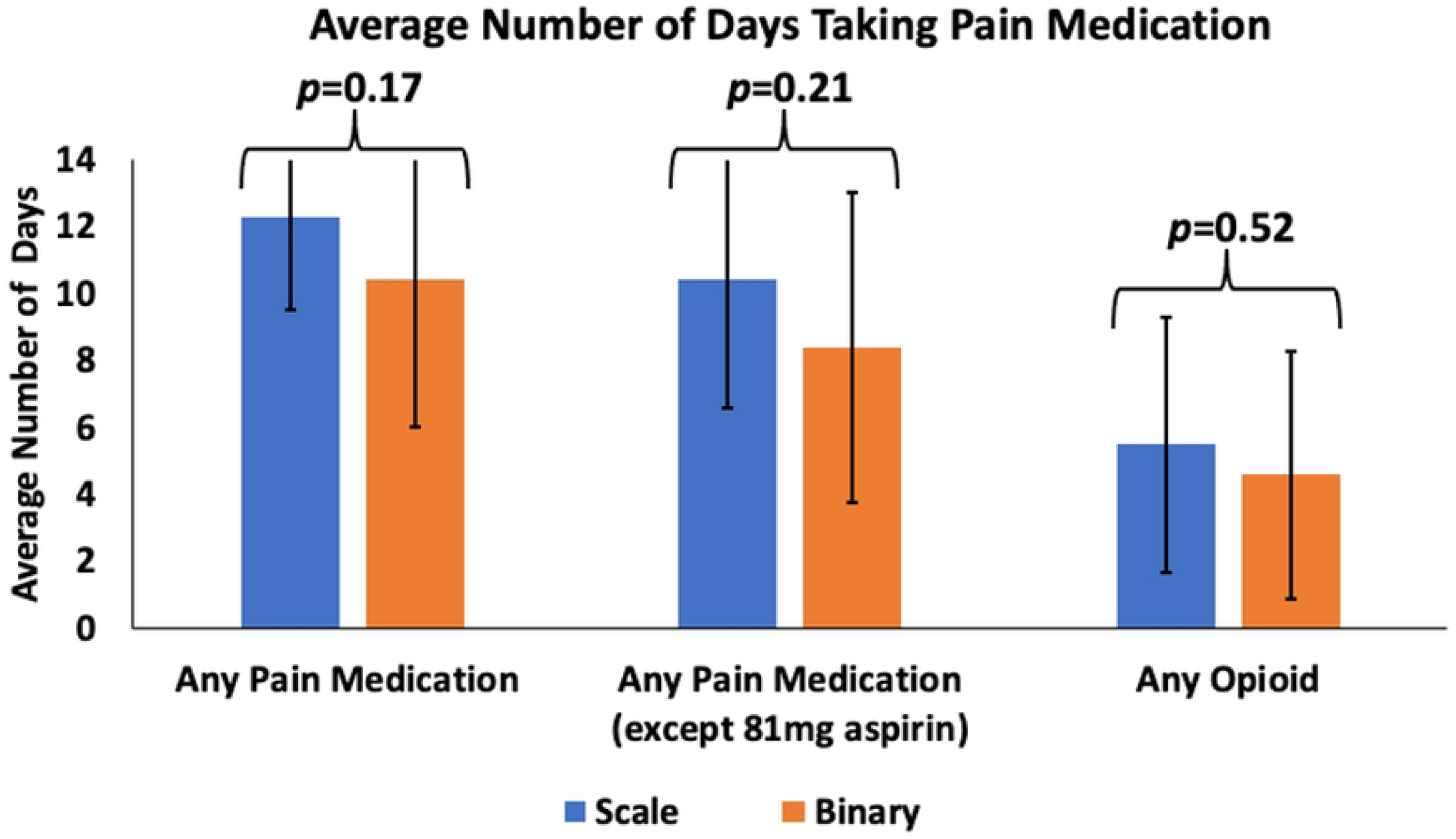
Average Number of Days Taking Pain Medications. For the 14 day post-operative period, patients were asked about the average number of days taking any pain medication, any pain medication excluding aspirin, and any opioid.

On POD 14, patients were asked about their satisfaction and side effects over the two weeks. Both groups reported high levels of patient satisfaction, with no statistically significant difference between the groups (8.0 ± 2.0 and 7.5 ± 3.4 out of 10 for the Scale and Binary groups, respectively; *p*=0.63, **Supplemental Table 4**). There was no significant difference between the average knee flexion for patients in both groups approximately two weeks after surgery (Scale (97° ±11°, Binary 89° ±19°; *p*=0.08, **Supplemental Table 4**). Over half of patients in both groups reported experiencing no side effects (60% in the Scale group and 57% in the Binary group), and no patients in either group reported urinary retention. Several patients in each group reported experiencing nausea, itching, decreased appetite, drowsiness, and headache (**Supplemental Table 5 and Supplemental Fig 1**).

## Discussion

This pilot prospective, randomized controlled study demonstrates that framing pain assessment in terms of tolerability rather than the traditional numerical scale could lead to reductions in pain medication use. Patients in the Scale group used any kind of pain medication for fewer days than patients in the Binary group, although this did not reach statistical significance (12.3 days versus 10.4 days, respectively, *p=*0.17). Patients in the Scale group consumed significantly more MMEs than the Binary group (*p*=0.03). There were high levels of patient satisfaction in both groups, with no significant difference between the experimental groups.

One possible explanation for our findings is that shifting the focus from a numerical value to tolerability may influence the patients’ perceptions of pain and thus influence their choice to take pain medication. Having patients assess their pain on a scale from 0 to 10 could inadvertently communicate to a patient that the goal should be for their pain to be 0, and therefore, patients could feel more encouraged to take pain medication. However, when patients are asked if their pain is tolerable versus intolerable, one message that might be delivered to the patient is that the goal is for the pain to be tolerable (and that the presence of pain is normal and expected). Despite the significant differences in how much pain medication was taken, there was no difference in the average degree of knee flexion between both groups.

The implication of these findings is that a dichotomous tolerability-based pain scale seems to be a practical alternative to the traditional numerical scale assessment in the clinical setting, with potential benefits associated with it. Across the United States, there are ongoing efforts to reduce the consumption of opioids, and implementation of a tolerability assessment of pain could be an important step towards better optimizing pain management and reducing opioid consumption. This pilot prospective clinical study could serve as the basis for further studies investigating whether tolerability-based assessments of pain could be more broadly applied.

There are several limitations to this paper that are important to address. Firstly, the limited sample size reduces the power of the study, thus making it challenging to generalize findings to a larger patient population. Additionally, patients in this study were mostly relatively young and otherwise healthy, and therefore, these findings may not be applicable to older patients with more comorbidities or patients undergoing other types of surgical procedures. Lastly, this study only followed patients for two weeks after surgery and, therefore, does not investigate the long-term effects, if any, of using an alternative pain assessment.

Future plans for this pilot trial include to expanding it to a larger group of enrolled patients at our center to potentially allow for more definitive conclusions to be drawn from the study. We will further attempt to include multiple other institutions and potentially other surgical interventions to increase the generalizability of these findings.

## Conclusions

This pilot prospective randomized clinical trial introduces the concept of using a dichotomous tolerability-based pain assessment in the post-operative period. Asking patients whether their pain is tolerable or intolerable instead of using the standard 10-point NPRS scale may lead to less opioid use in the early postoperative period.

## Data Availability

All relevant data are within the manuscript and its Supporting Information files

## Supporting Information

**Supplemental Figure 1:**
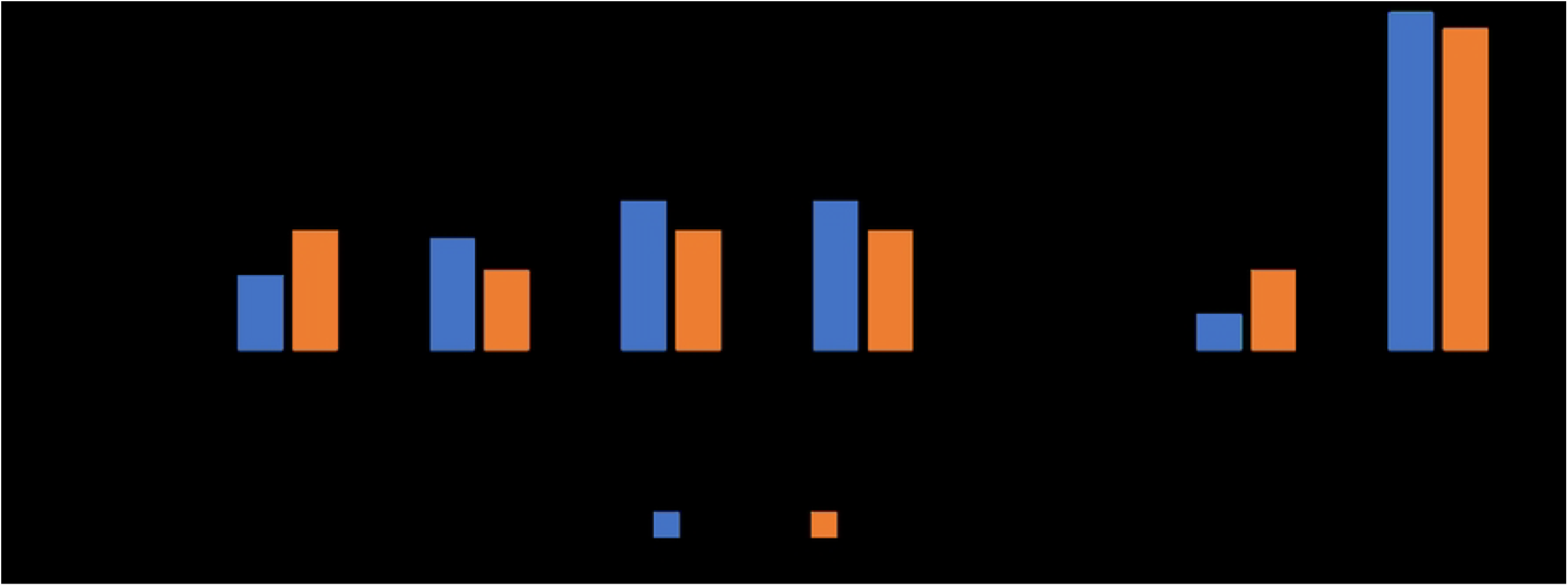
Side Effects Before the First Follow-Up Appointment.

**Supplemental Table 1: Distribution of Allografts and Autografts Between Groups**. Data is presented as number (percentage). P-value was calculated using a Chi-squared test.

**Supplemental Table 2: Average Number of Days Taking Pain Medications in the Post-Operative Period**. Data is presented as average ± standard deviation.

**Supplemental Table 3: Average Doses per Day of Opioids in the Post-Operative Period**.

Data is presented as average ± standard deviation.

**Supplemental Table 4: Overall Satisfaction with Pain Management**. All participants were asked to rate their satisfaction with their pain medication on a scale from 0 to 10, with 0 being the least satisfied and 10 being the most satisfied. Data is presented as average ± standard deviation.

**Supplemental Table 5: Side Effects Experienced Before First Follow-Up Visit**.

